# The impact of vaccination on *Neisseria gonorrhoeae* antimicrobial resistance and prevalence in men who have sex with men: a mathematical modelling study

**DOI:** 10.1101/2020.09.14.20192062

**Authors:** Janneke C.M. Heijne, Maria Xiridou, Katy M.E. Turner, Maartje Basten, Maartje Visser, Birgit van Benthem, Nicola Low

## Abstract

**Background:** *Neisseria gonorrhoeae* (gonorrhoea) and antimicrobial-resistant (AMR) gonorrhoea infections disproportionately affect men who have sex with men (MSM). Vaccine development is challenging, but a *N. meningitidis* (group B) vaccine given to children and young adults was associated with a ca. ~30% reduction in gonorrhoea diagnoses. We investigated the impact of vaccination on *N. gonorrhoeae* AMR development and transmission in MSM.

**Methods:** We developed a compartmental model of *N. gonorrhoeae* transmission among MSM. AMR to ceftriaxone was incorporated as a stepwise increases in minimum inhibitory concentrations and eventual resistance (MIC drift). We estimated the impact of a partially protective vaccine (reducing susceptibility; 2-years protection) targeting high sexual activity MSM on AMR and prevalence until 2050. We performed sensitivity analyses assuming different levels of vaccine effectiveness (VE) and other modes of vaccine action.

**Findings:** Gonorrhoea model prevalence was 3·4% (95% credible interval 3·2% – 3·8%) in all MSM, 12·5% (95% credible interval 12·1% – 12·7%) in high sexual activity MSM. A vaccine with 30% VE cannot prevent AMR, even with high uptake or durable protection. However, it increases time to AMR development by several years. For a fixed uptake of 40% a vaccine needs a minimum VE of 90% to prevent AMR development completely. A vaccine providing complete protection to infection for those vaccinated was most effective in reducing population prevalence and preventing AMR.

**Interpretation:** A vaccine that has limited efficacy for the prevention of gonorrhoea could delay AMR development in MSM, providing time for developing new antimicrobials and more efficacious vaccines.

**Funding:** None

## Introduction

Sexually transmitted *Neisseria gonorrhoeae* (gonorrhoea) is an urgent public health concern because of the threat of antimicrobial resistance (AMR), according to the World Health Organizatio.^1^ In many countries, high proportions of tested *N. gonorrhoeae* isolates are resistant to all previously used classes of antimicrobials. Ceftriaxone, often in combination with azithromycin, is the only recommended first-line treatment regimen, but extensively drug resistant strains are being increasingly reported.^2^ Whilst the precise mechanisms of development of *N. gonorrhoeae* resistance to ceftriaxone are incompletely understood and probably vary geographically, the steady shift towards higher values in the peak of the distribution of minimum inhibitory concentrations (MIC)^3,4^ indicates a drift towards resistance. Thus far, strains with resistance to ceftriaxone, defined by the European Union Committee on Antimicrobial Susceptibility Testing as MIC >0·125 mg/L,^5^ have not been detected in the Dutch Gonococcal Resistance to Antimicrobials Surveillance (GRAS) program. However, higher proportions of isolates with reduced susceptibility (MIC range) were seen in 2019 compared with previous years and two isolates reached the threshold value for resistance in 2019.^6^

Vaccination is an intervention that could both reduce the overall incidence of gonorrhoea and the emergence of resistance in *N. gonorrhoeae*.^7-9^ Interest in gonorrhoea vaccine development has increased since the publication of a study from New Zealand about a vaccine, based on the outer membrane vesicle of *N. meningitidis* group B (MeNZB). The vaccine was given to children and young adults to prevent meningococcal disease and was associated with short lasting (2-3 years) reductions of 31% (95% CI 21 – 39%) in gonorrhoea incidence^10^ and 24% (95% CI 1 – 42%) in hospitalizations caused by gonorrhoea.^11^ Even a partially protective vaccine might have public health benefits amongst groups at disproportionate risk of gonorrhoea and of AMR, such as men who have sex with men (MSM).^12,13^ At Dutch sexual health centers 77% of gonorrhoea diagnoses were among MSM in 2019.^6^ Gonorrhoea diagnosis rates among MSM have been increasing in many countries.^6,14,15^ Reasons for this increase might include reduced condom use amongst MSM with HIV infection who are taking antiretroviral treatment as prevention (TasP)^16^ or those without HIV infection taking pre-exposure prophylaxis (PrEP),^17^ and the message that undetectable (HIV viral load) equals untransmissible (U=U).^18^ The spread of untreated antimicrobial-resistant *N. gonorrhoeae* strains could also increase the incidence of gonorrhoea.^19-23^

Mathematical modelling studies of the effects of a gonorrhoea vaccine on AMR in MSM have focused on the impact of the introduction of a single resistant gonorrhoea strain on AMR persistence.^24,25^ These studies found that vaccination greatly reduces the probability of an outbreak with a resistant strain, and that, when vaccine protection is long enough, the incidence of gonorrhoea can be greatly reduced. Studies of the general impact of gonorrhoea vaccination on transmission have found a substantial reduction in the prevalence of infection.^26,27^ However, modelling studies have not investigated how vaccination affects the development of *N. gonorrhoeae* resistance to ceftriaxone within a population and its subsequent transmission. The objectives of this study were to investigate the impact of a gonorrhoea vaccine on the development of *N. gonorrhoeae* AMR to ceftriaxone and on transmission among MSM.

## METHODS

We developed a deterministic model that describes the development of AMR to ceftriaxone monotherapy, the first-line treatment for gonorrhoea in the Netherlands since 2006,^28,29^ *N. gonorrhoeae* transmission among MSM and *N. gonorrhoeae* testing and treatment practices at sexual health centers in the Netherlands. The data sources and model are described below. The model equations are described in Supplement 1.

### Sexual behaviour

MSM enter the model when they become sexually active, and leave after 45 years (Table 1). Upon entry in the model, they are divided into three activity classes (low, medium, high), which differ in sexual partner change rates (Supplement 2). In the model, men can change their sexual behaviour throughout their sexually active life. The redistribution is proportional to the size of the activity class, meaning that men in the larger sexual activity classes are less likely to change to other classes than men from smaller sexual activity classes.^30^ Mixing between sexual activity classes is incorporated in one parameter that can range from assortative to proportionate mixing. The partner change rates and percentages of people in each activity class are informed by the Amsterdam Cohort Studies on HIV and AIDS, an open cohort of MSM ^S31,S32^ (Supplement 2).

**Table 1.**
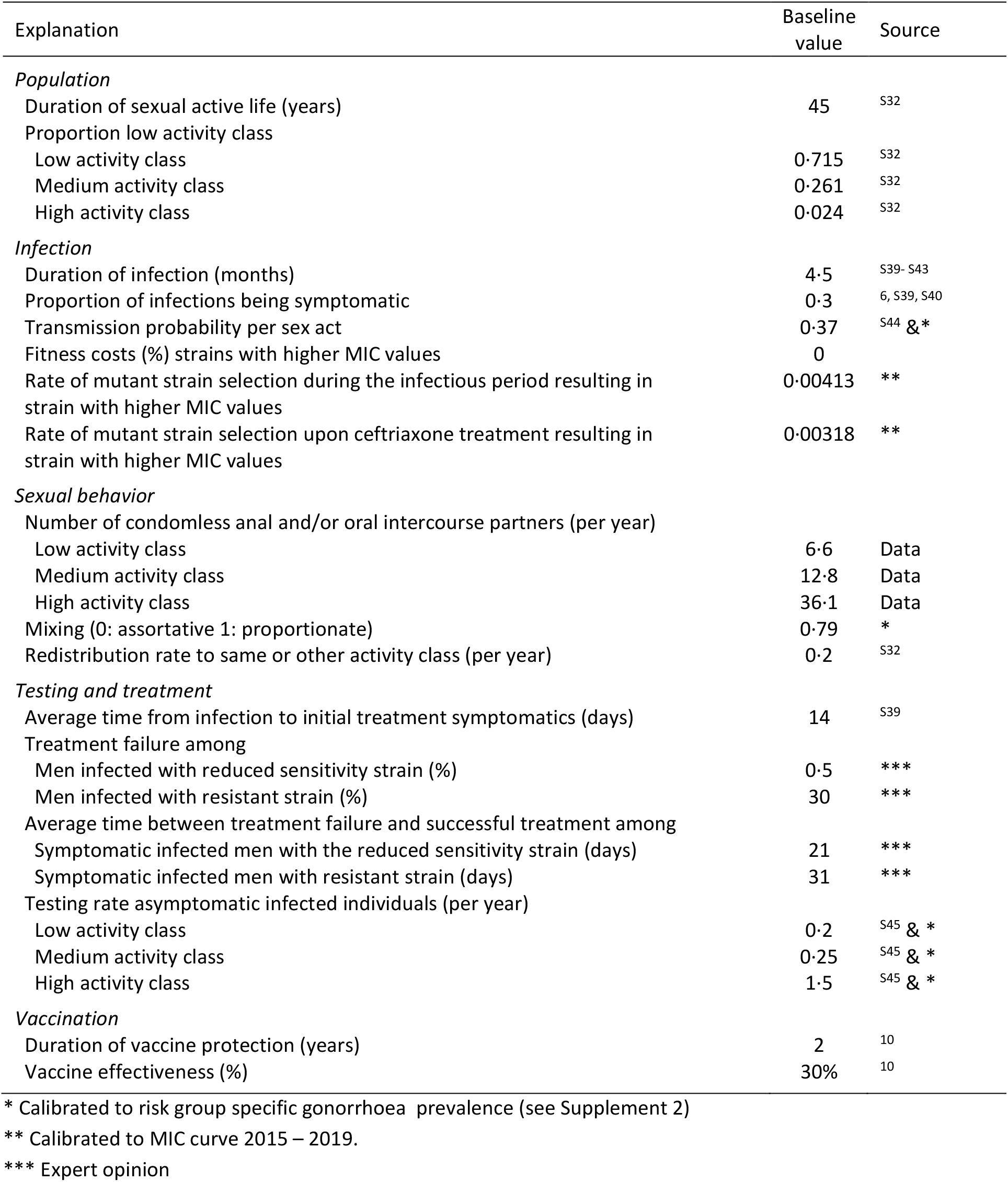
Baseline parameters of the model and their values.

### *N. gonorrhoeae* infection and emergence of resistance

In the model, MSM can either be susceptible to *N. gonorrhoeae* infection or infected (Figure 1 and Supplementary Figure S1.1). For simplicity, we consider the urogenital, anorectal and pharyngeal sites of infection together. Upon infection, there is a probability of developing symptoms. The probability of developing symptoms vary across anatomical locations, with urogenital infections being mostly symptomatic, whereas anorectal and pharyngeal infections being mostly asymptomatic.^6^ Since about 30% of all infected MSM visiting Dutch sexual health centers have a urogenital gonorrhoea infection (either single or co-infected at other locations),^6^ we assumed the percentage symptomatic to be 30%. At any time, men can clear the infection naturally.

**Figure 1.**
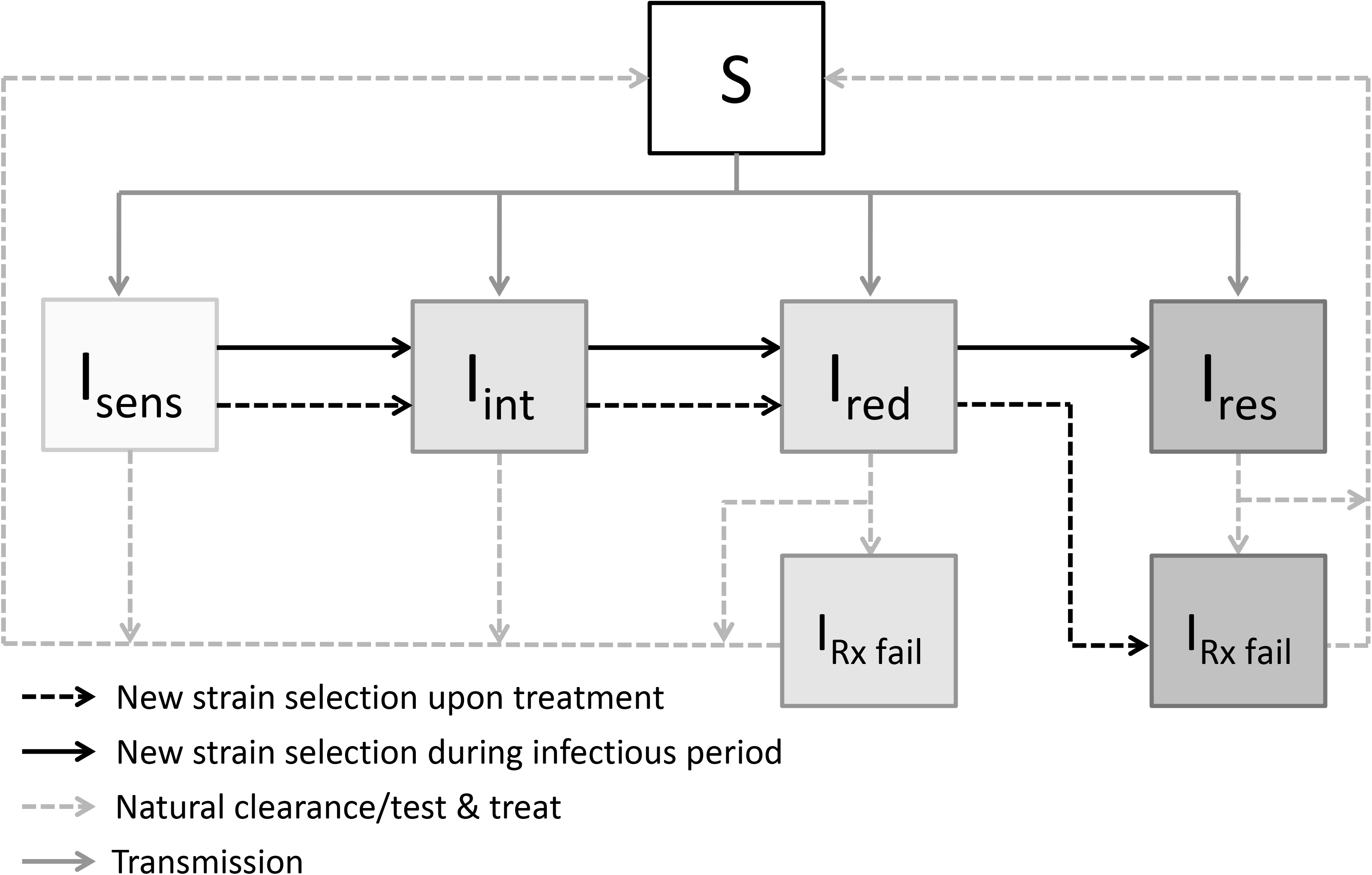
Schematic overview of multistage gonorrhoea AMR development in the model in symptomatically infected MSM. People can either be susceptible (S) or infected with strains defined as: fully sensitive to ceftriaxone (I_sens_). intermediate sensitivity (I_int_). reduced sensitivity (I_red_) or resistant (I_res_). The I_RX fail_ compartment denotes ceftriaxone treatment failure, but successful alternative treatment. Please note that this overview is the same for each risk class. A full schematic overview of the model can be found in Supplemental material 1.

Development of *N. gonorrhoeae* AMR to ceftriaxone is incorporated as a multistage unidirectional process, assuming that the drift in MIC distribution over time could result from the accumulation of genetic mutations that increase the MIC and eventually result in resistance.^5^ For the model, we defined four groups of strains, according to categories of MIC: fully sensitive (MIC <=0·016 mg/L), intermediate sensitivity (MIC >0·016 and <=0·064 mg/L), reduced sensitivity (MIC >0·064 and <=0·125 mg/L) and resistant (MIC values >0·125 mg/L). We use the term 'sensitivity' to antimicrobials to distinguish from the susceptibility of MSM to *N. gonorrhoeae* infection. Due to lack of data, we assumed that there is no fitness cost for each group of strains. However, we performed an uncertainty analysis in which we assumed that the resistant strain has a 5% lower transmission probability.

The exact mechanism of the drift in MIC distribution is unknown. In the model, we distinguish two different rates. The first assumes selection for mutant strains upon treatment by exposure of *N. gonorrhoeae* to ceftriaxone (further referred to as new strain selection upon ceftriaxone treatment). The second assumes selection for mutant strains during the infectious period through exposure of *N. gonorrhoeae* to antimicrobials given for other infections^23^ (further referred to as new strain selection during infectious period). This process is independent of *N. gonorrhoeae* testing and treatment. In the baseline analyses we include both rates, but perform sensitivity analyses assuming one or the other rate.

### Testing and follow-up for *N. gonorrhoeae*

In the model, testing of MSM with symptoms is independent of sexual activity class. Testing of MSM without symptoms does depend on the activity class, where MSM in the highest activity class visit the sexual health centers most frequently and are most likely to be tested (Table 1). It is assumed that everyone tested positive receives treatment.

Treatment is incorporated in the model with four assumptions (Figure 1). First, everyone who is tested positive initially receives ceftriaxone as empirical first line treatment, as recommended in Dutch sexual health centers. Testing for sensitivity to antimicrobials is routinely performed for surveillance purposes and only determines treatment decisions if patients return with persistent symptoms. Second, the dose of ceftriaxone will be sufficient to clear the *N. gonorrhoeae* strain with reduced sensitivity in all but a very small proportion, since ceftriaxone treatment failure has been documented with MIC values lower than 0·125 mg/L.^21,22^ Third, of the men infected with a resistant strain and treated with ceftriaxone, most will still clear the infection but the fraction of men with treatment failure will be higher than for those infected with the reduced sensitivity strain. Fourth, upon treatment for MSM infected with the fully, intermediate or reduced sensitivity strain, selection for mutant strains could happen (process one as described above). This results in moving to the next compartment with increased MIC levels.

MSM with symptomatic gonorrhoea who fail treatment (either because they are already infected with a reduced sensitivity or resistant strain or they develop resistance upon treatment) are assumed to have persistent symptoms and return to the clinic. They receive gonorrhoea culture and antimicrobial susceptibility testing (if not already done), followed by effective treatment with alternative antibiotics. The time between testing and effective treatment is assumed to be longer for men infected with a resistant strain than with a strain with reduced sensitivity. The asymptomatically infected MSM with treatment failure do not return to the clinic because a test of cure after treatment for asymptomatic gonorrhoea is not the standard of care in the Netherlands. These men may transmit the strain with reduced sensitivity or the resistant one.

### Vaccination

In the model, vaccination uptake depends on the sexual activity class. For simplicity, we assumed that only MSM susceptible to gonorrhoea receive the vaccine. The vaccine is assumed to provide short lasting protection, and MSM can be vaccinated multiple times. Because the MeNZB vaccine is not directed against gonococcal antigens, we assumed that the modelled vaccine provides the same level of partial protection to all vaccinated individuals (a leaky vaccine). In the main analyses, we assumed that the vaccine reduces susceptibility to infection. The exact mode of vaccine action is unknown, so we performed uncertainty analyses assuming two alternative modes: that the partially protective vaccine reduced the transmissibility or the duration of infection. We also performed an uncertainty analysis assuming that the future vaccine is an all-or-nothing vaccine, i.e. providing either complete or no protection to infection for those vaccinated.

### Model calibration

Details of the calibration procedure can be found in the Supplemental material, sections 2 and 3. In short, the unknown parameters that were unrelated to AMR development (mixing parameter, transmission probability per sex act and the activity class specific testing uptake) were calibrated to the activity class-specific percentage of gonorrhoea (pharyngeal/anorectal/urogenital) diagnosed in MSM in the Amsterdam Cohort Studies. The first 500 parameter sets that had the smallest sum of the squared residuals were used to further calibrate the model to reflect the current AMR situation in the Netherlands. To do this, the model was run into steady state with a fully sensitive strain of *N. gonorrhoeae* to ceftriaxone, after which ceftriaxone treatment was introduced in 2006. For each parameter set, the two mutant selection rates (upon treatment and during infectious period) were obtained by fitting the model to the MIC curve for ceftriaxone from the Dutch GRAS program for the years 2015 – 2019.^6^ This resulted in 343 useful mutant selection rates that were used throughout the analyses.

### Analyses

In all analyses, vaccination was introduced in 2021 (15 years after the introduction of ceftriaxone). The effects of vaccination were obtained from model outputs until 2050. In the main analyses, only high risk MSM are targeted for vaccination and the input value for vaccine effectiveness (VE) is 30%, with uncertainty analyses for other levels of VE. Results are presented as the overall prevalence of gonorrhoea, prevalence of the non-resistant strains (fully sensitive, intermediate and reduced sensitivity) and prevalence of the resistant strain. For readability, the best fitted parameter set was used in the figures, and all other results are provided as medians and 95% credible intervals (95% CrI) form all fitted parameter sets.

## RESULTS

In the absence of vaccination, the overall gonorrhoea prevalence in MSM was 3·4% (95% Cr 3·2% – 3·8%), with a prevalence of 12·5% (95% Cr 12·1% – 12·7%) in high activity MSM, 5·0% (95% Cr 4·5% – 5·4%) in medium and 2-5% (95% Cr 2·4% – 3·0%) in low activity MSM (Figure 2A). The prevalence in high and medium activity MSM is similar to the Amsterdam Cohort Studies data, but the prevalence in the low risk groups is slightly higher than the data (Table S3.2 in Supplement). The distribution of MICs for the years 2015 – 2019 from the model output was comparable to the GRAS data (Figure 2B). In the model, AMR developed slowly and it would take multiple years for the prevalence of the resistant strain to reach the 5% threshold value for switching antibiotics, as recommended by WHO (Figure 2C).

**Figure 2.**
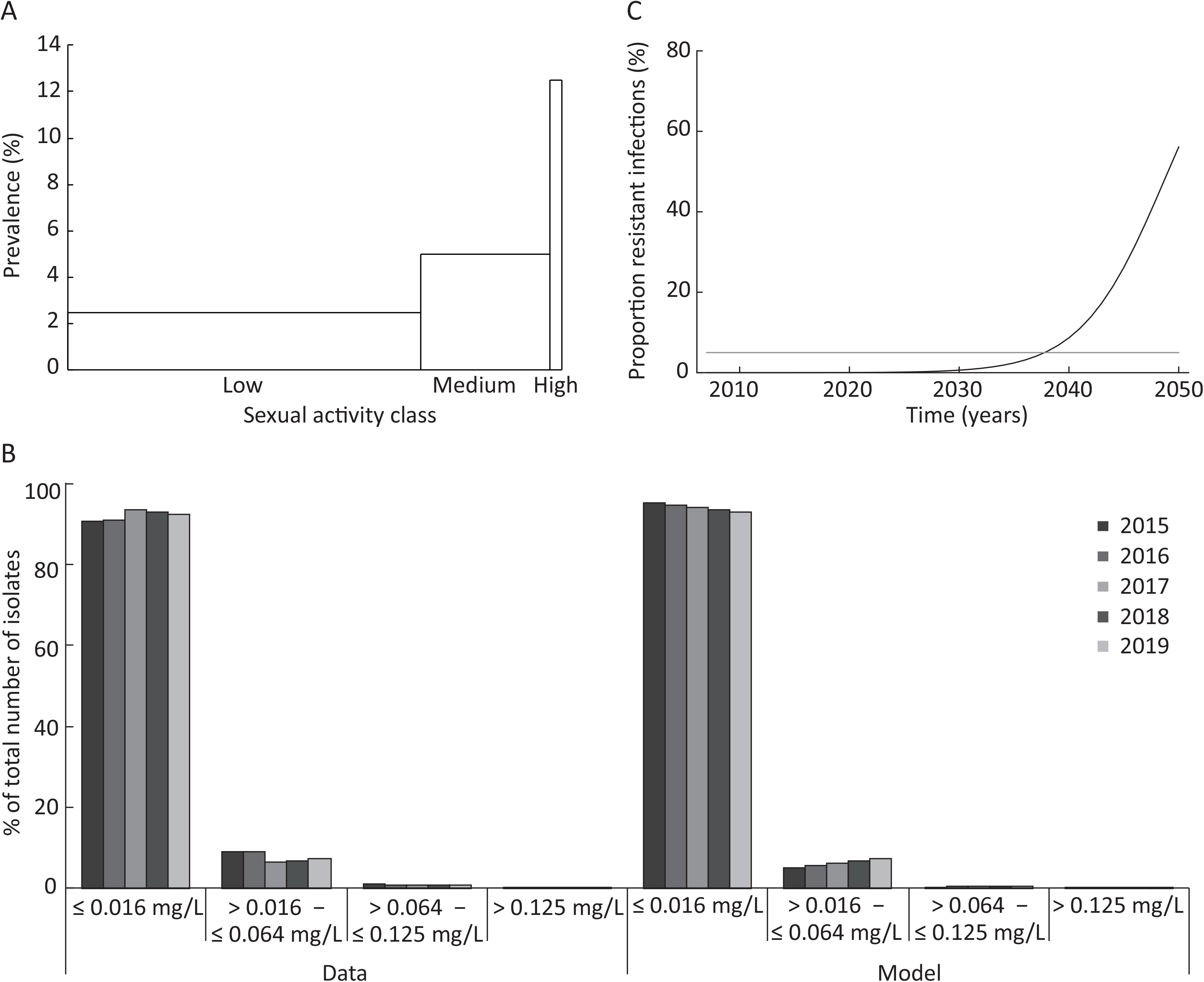
Gonorrhoea prevalence and AMR in the absence of vaccination. (A) Baseline gonorrhoea prevalence in the three sexual activity classes; the width of the bars are proportional to the proportion in the sexual activity class; (B) MIC curve for ceftriaxone from AMR surveillance data^6^ and model output; (C) Proportion of resistant infections (MIC>0-125 mg/L); the grey line denotes the 5% threshold value recommended by WHO for switching antibiotics.

### Effects of vaccination

Vaccinating high activity MSM with a partially protective vaccine that reduces susceptibility to *N. gonorrhoeae* by 30% and immunity lasting 2 years cannot prevent the development of a fully resistant strain, even at high vaccine uptake (Figure 3A-C). However, vaccination delays the development of AMR at all levels of vaccine uptake. On average, every 10% increase in coverage confers a gain of half a year before AMR prevalence takes off. For a fixed vaccine uptake of 40% of high risk MSM per year, a partially protective vaccine needs to have a VE of at least 90% to prevent AMR and eliminate gonorrhoea (Figure 3D-F).

**Figure 3.**
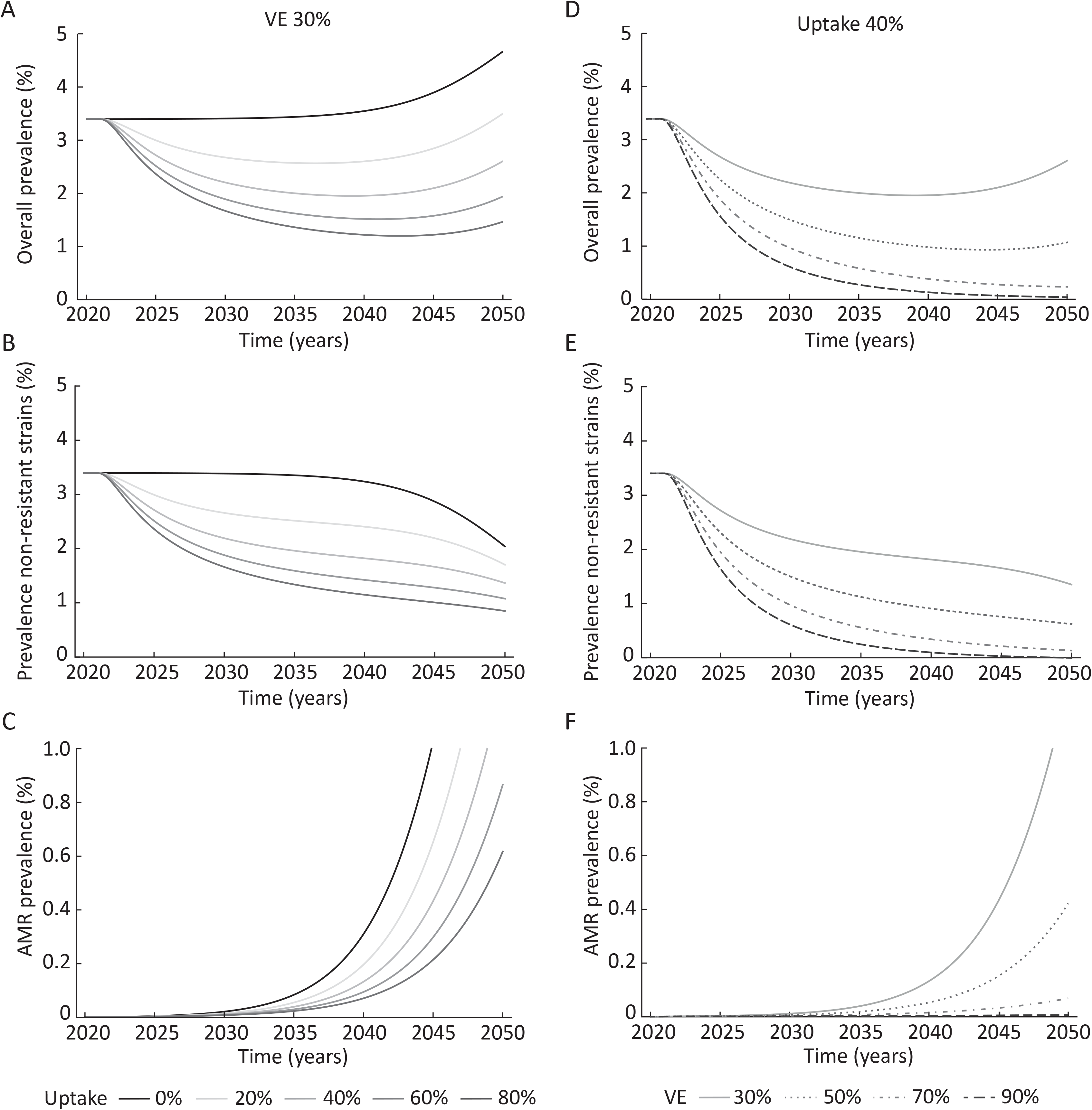
Effect on prevalence of a partially protective vaccine that reduces susceptibility to *N. gonorrhoeae* for a vaccine with a fixed vaccine effectiveness (VE) of 30% and different vaccination uptake in high activity MSM (A-C) or a fixed vaccination uptake of 40% and different VE (D-F). Vaccination is introduced in 2021.

When looking at the effects of vaccination on overall prevalence, a vaccine with limited VE is able to reduce the overall prevalence initially, after which it increases again. This is a result of two opposing forces. Vaccination reduces the prevalence of the non-resistant strains. However, this decrease is not enough to prevent the development of AMR. Since the resistant strain takes longer to treat or fails treatment more often, AMR prevalence increases, and the overall prevalence increases again (Figure 3A-C). For high VE, gonorrhoea is eliminated before AMR can develop, and hence the overall prevalence decreases (Figure 3D-F).

A longer duration of vaccine protection does not prevent AMR when VE is 30% (Table 2). However, increasing vaccination uptake by also targeting MSM in the medium activity class does prevent AMR when uptake is 40% or higher when VE is 30%. For higher VE (>50%) AMR can be prevented if the duration of vaccine protection is 5 years (Table 3). If the resistant strain is 5% less fit than the non-resistant strains, AMR transmission is slower, so vaccination (even with a low VE) is effective in preventing AMR compared to no fitness costs.

**Table 2.**
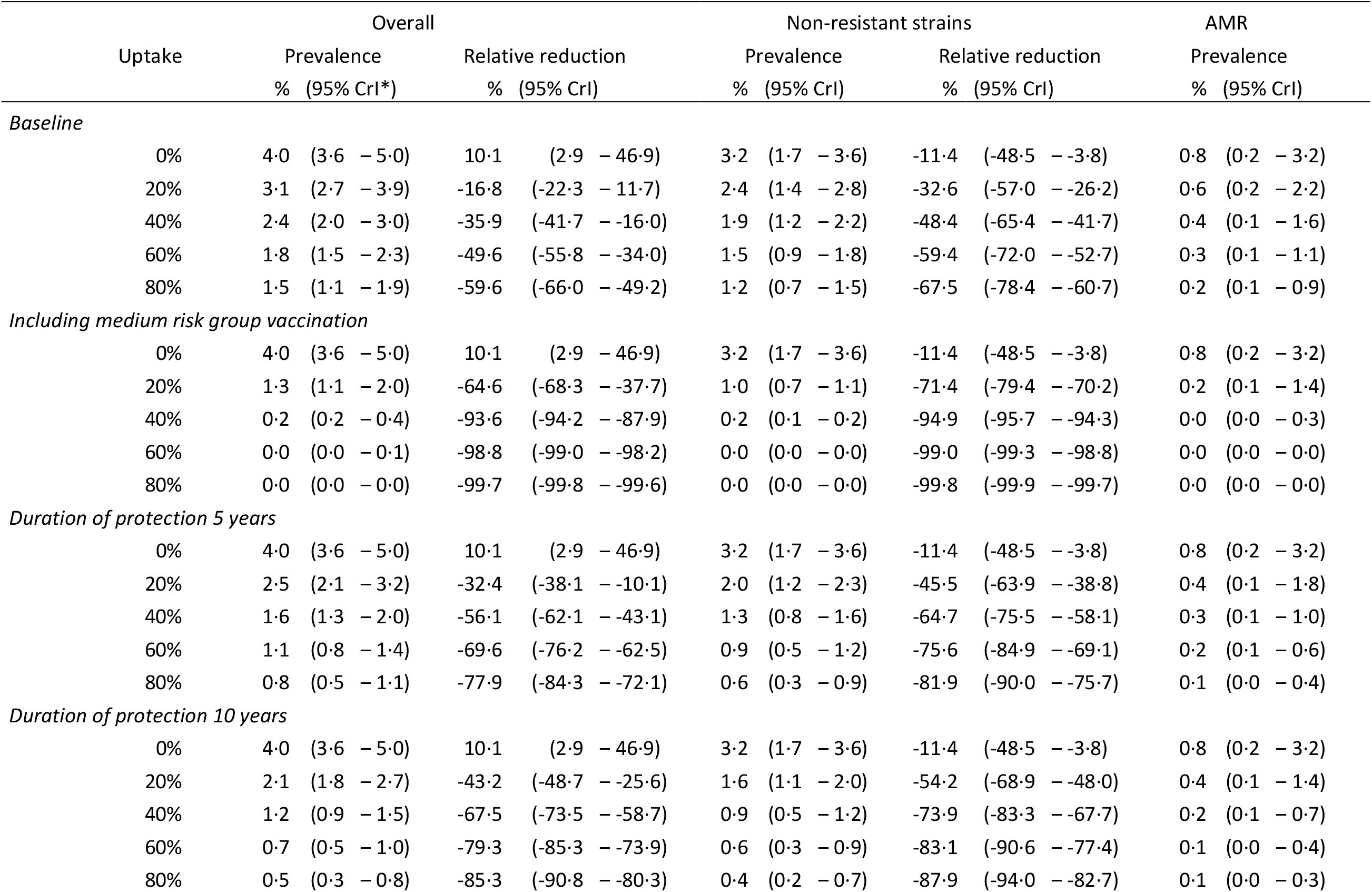
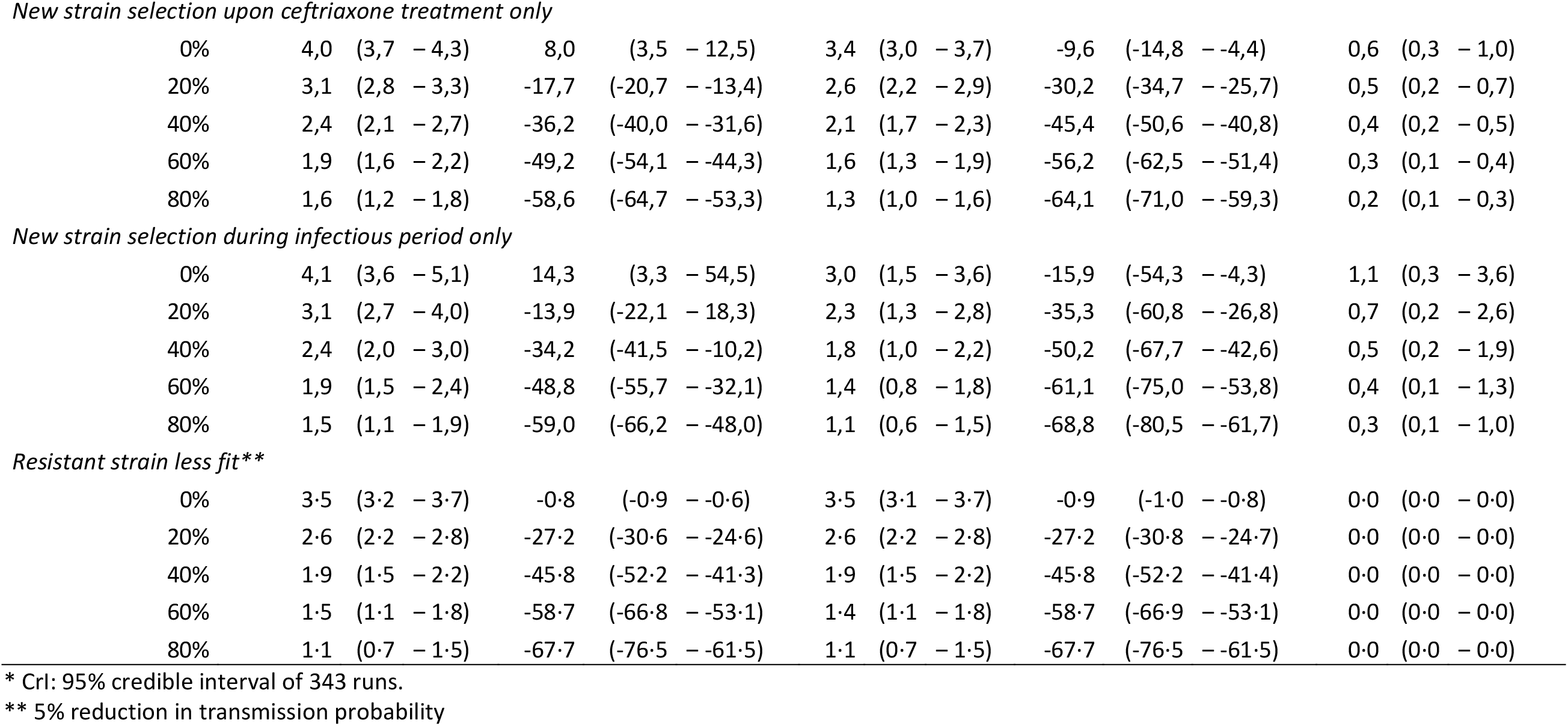
Uncertainty analyses on the impact of a partially protective vaccine that reduces susceptibility to *N. gonorrhoeae* for a vaccine with a fixed vaccine effectiveness of 30% and different vaccination uptake on prevalence in 2050.

**Table 3.**
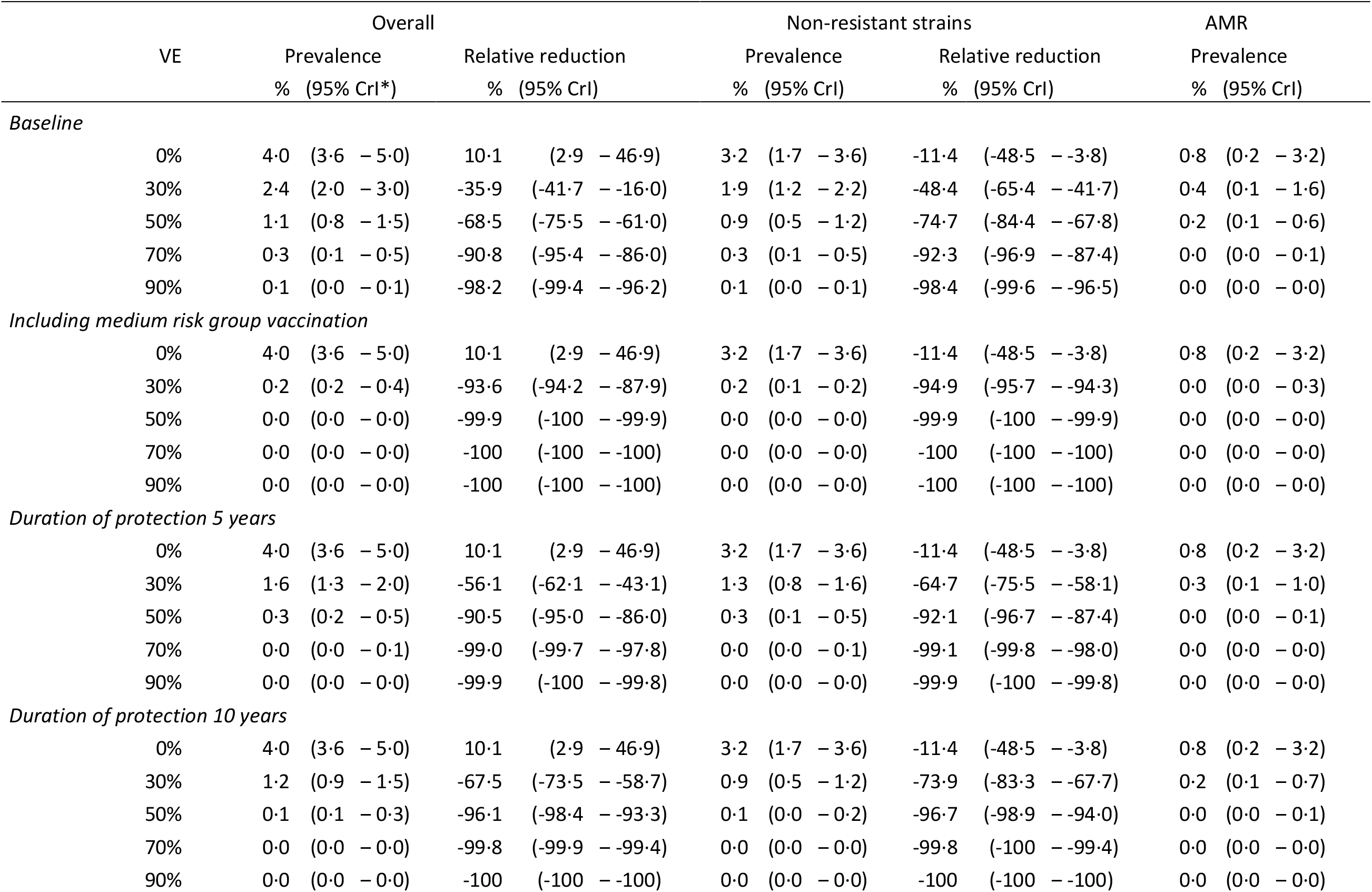
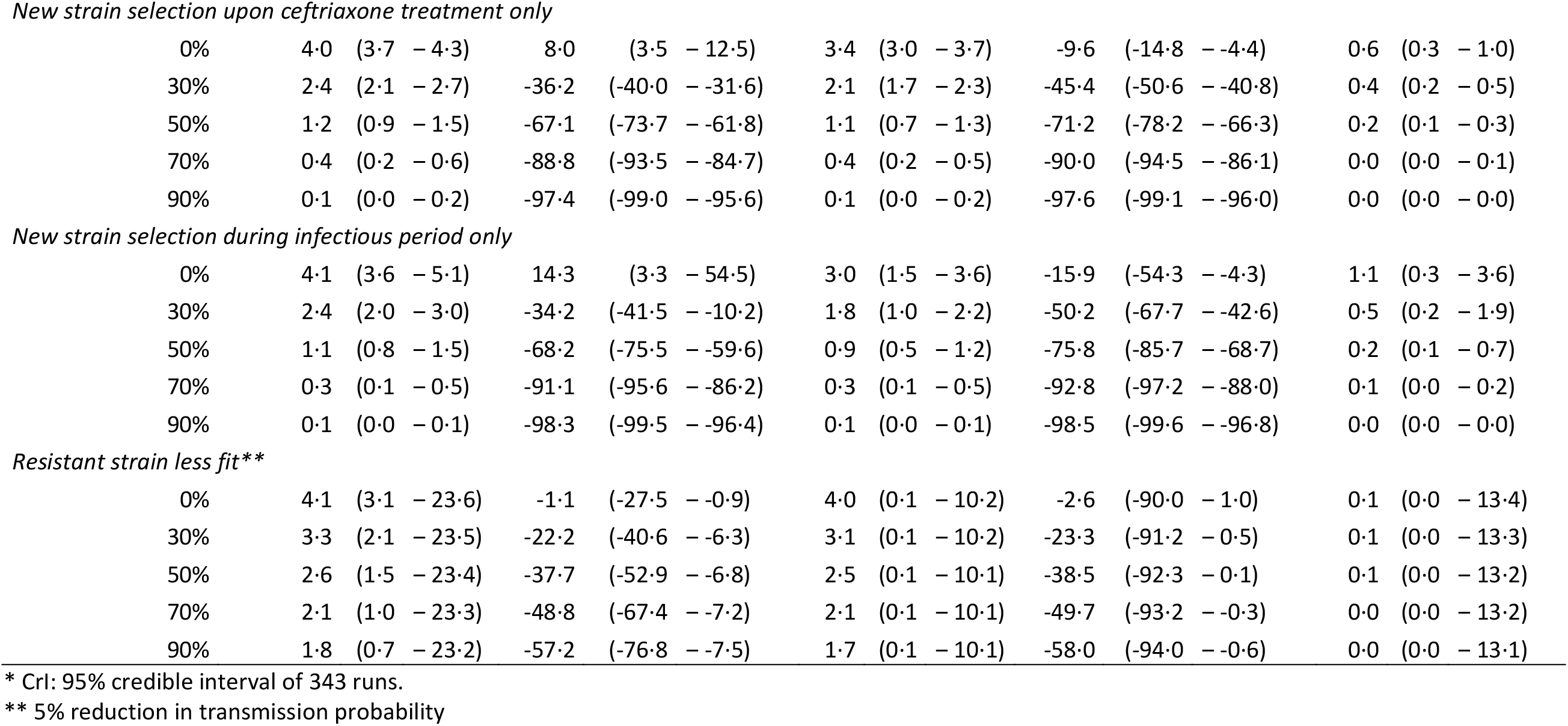
Uncertainty analyses on the impact of a partially protective vaccine that reduces susceptibility to *N. gonorrhoeae* for a vaccine with a fixed vaccine uptake of 40% and different vaccine effectiveness (VE) on prevalence in 2050.

|  | Overall |  |  |  | Non-resistant strains |  |  |  | AMR |  |
| --- | --- | --- | --- | --- | --- | --- | --- | --- | --- | --- |
| VE | Prevalence |  | Relative reduction |  | Prevalence |  | Relative reduction |  | Prevalence |  |
|  | % | (95% CrI*) | % | (95% CrI) | % | (95% CrI) | % | (95% CrI) | % | (95% CrI) |
| *Baseline* |  |  |  |  |  |  |  |  |  |  |
| 0% | 4.0 | (3.6 – 5.0) | 10.1 | (2.9 – 46.9) | 3.2 | (1.7 – 3.6) | -11.4 | (-48.5 – -3.8) | 0.8 | (0.2 – 3.2) |
| 30% | 2.4 | (2.0 – 3.0) | -35.9 | (-41.7 – -16.0) | 1.9 | (1.2 – 2.2) | -48.4 | (-65.4 – -41.7) | 0.4 | (0.1 – 1.6) |
| 50% | 1.1 | (0.8 – 1.5) | -68.5 | (-75.5 – -61.0) | 0.9 | (0.5 – 1.2) | -74.7 | (-84.4 – -67.8) | 0.2 | (0.1 – 0.6) |
| 70% | 0.3 | (0.1 – 0.5) | -90.8 | (-95.4 – -86.0) | 0.3 | (0.1 – 0.5) | -92.3 | (-96.9 – -87.4) | 0.0 | (0.0 – 0.1) |
| 90% | 0.1 | (0.0 – 0.1) | -98.2 | (-99.4 – -96.2) | 0.1 | (0.0 – 0.1) | -98.4 | (-99.6 – -96.5) | 0.0 | (0.0 – 0.0) |
| *Including medium risk group vaccination* |  |  |  |  |  |  |  |  |  |  |
| 0% | 4.0 | (3.6 – 5.0) | 10.1 | (2.9 – 46.9) | 3.2 | (1.7 – 3.6) | -11.4 | (-48.5 – -3.8) | 0.8 | (0.2 – 3.2) |
| 30% | 0.2 | (0.2 – 0.4) | -93.6 | (-94.2 – -87.9) | 0.2 | (0.1 – 0.2) | -94.9 | (-95.7 – -94.3) | 0.0 | (0.0 – 0.3) |
| 50% | 0.0 | (0.0 – 0.0) | -99.9 | (-100 – -99.9) | 0.0 | (0.0 – 0.0) | -99.9 | (-100 – -99.9) | 0.0 | (0.0 – 0.0) |
| 70% | 0.0 | (0.0 – 0.0) | -100 | (-100 – -100) | 0.0 | (0.0 – 0.0) | -100 | (-100 – -100) | 0.0 | (0.0 – 0.0) |
| 90% | 0.0 | (0.0 – 0.0) | -100 | (-100 – -100) | 0.0 | (0.0 – 0.0) | -100 | (-100 – -100) | 0.0 | (0.0 – 0.0) |
| *Duration of protection 5 years* |  |  |  |  |  |  |  |  |  |  |
| 0% | 4.0 | (3.6 – 5.0) | 10.1 | (2.9 – 46.9) | 3.2 | (1.7 – 3.6) | -11.4 | (-48.5 – -3.8) | 0.8 | (0.2 – 3.2) |
| 30% | 1.6 | (1.3 – 2.0) | -56.1 | (-62.1 – -43.1) | 1.3 | (0.8 – 1.6) | -64.7 | (-75.5 – -58.1) | 0.3 | (0.1 – 1.0) |
| 50% | 0.3 | (0.2 – 0.5) | -90.5 | (-95.0 – -86.0) | 0.3 | (0.1 – 0.5) | -92.1 | (-96.7 – -87.4) | 0.0 | (0.0 – 0.1) |
| 70% | 0.0 | (0.0 – 0.1) | -99.0 | (-99.7 – -97.8) | 0.0 | (0.0 – 0.1) | -99.1 | (-99.8 – -98.0) | 0.0 | (0.0 – 0.0) |
| 90% | 0.0 | (0.0 – 0.0) | -99.9 | (-100 – -99.8) | 0.0 | (0.0 – 0.0) | -99.9 | (-100 – -99.8) | 0.0 | (0.0 – 0.0) |
| *Duration of protection 10 years* |  |  |  |  |  |  |  |  |  |  |
| 0% | 4.0 | (3.6 – 5.0) | 10.1 | (2.9 – 46.9) | 3.2 | (1.7 – 3.6) | -11.4 | (-48.5 – -3.8) | 0.8 | (0.2 – 3.2) |
| 30% | 1.2 | (0.9 – 1.5) | -67.5 | (-73.5 – -58.7) | 0.9 | (0.5 – 1.2) | -73.9 | (-83.3 – -67.7) | 0.2 | (0.1 – 0.7) |
| 50% | 0.1 | (0.1 – 0.3) | -96.1 | (-98.4 – -93.3) | 0.1 | (0.0 – 0.2) | -96.7 | (-98.9 – -94.0) | 0.0 | (0.0 – 0.1) |
| 70% | 0.0 | (0.0 – 0.0) | -99.8 | (-99.9 – -99.4) | 0.0 | (0.0 – 0.0) | -99.8 | (-100 – -99.4) | 0.0 | (0.0 – 0.0) |
| 90% | 0.0 | (0.0 – 0.0) | -100 | (-100 – -100) | 0.0 | (0.0 – 0.0) | -100 | (-100 – -100) | 0.0 | (0.0 – 0.0) |
| *New strain selection upon ceftriaxone treatment only* |  |  |  |  |  |  |  |  |  |  |
| 0% | 4.0 | (3.7 – 4.3) | 8.0 | (3.5 – 12.5) | 3.4 | (3.0 – 3.7) | -9.6 | (-14.8 – -4.4) | 0.6 | (0.3 – 1.0) |
| 30% | 2.4 | (2.1 – 2.7) | -36.2 | (-40.0 – -31.6) | 2.1 | (1.7 – 2.3) | -45.4 | (-50.6 – -40.8) | 0.4 | (0.2 – 0.5) |
| 50% | 1.2 | (0.9 – 1.5) | -67.1 | (-73.7 – -61.8) | 1.1 | (0.7 – 1.3) | -71.2 | (-78.2 – -66.3) | 0.2 | (0.1 – 0.3) |
| 70% | 0.4 | (0.2 – 0.6) | -88.8 | (-93.5 – -84.7) | 0.4 | (0.2 – 0.5) | -90.0 | (-94.5 – -86.1) | 0.0 | (0.0 – 0.1) |
| 90% | 0.1 | (0.0 – 0.2) | -97.4 | (-99.0 – -95.6) | 0.1 | (0.0 – 0.2) | -97.6 | (-99.1 – -96.0) | 0.0 | (0.0 – 0.0) |
| *New strain selection during infectious period only* |  |  |  |  |  |  |  |  |  |  |
| 0% | 4.1 | (3.6 – 5.1) | 14.3 | (3.3 – 54.5) | 3.0 | (1.5 – 3.6) | -15.9 | (-54.3 – -4.3) | 1.1 | (0.3 – 3.6) |
| 30% | 2.4 | (2.0 – 3.0) | -34.2 | (-41.5 – -10.2) | 1.8 | (1.0 – 2.2) | -50.2 | (-67.7 – -42.6) | 0.5 | (0.2 – 1.9) |
| 50% | 1.1 | (0.8 – 1.5) | -68.2 | (-75.5 – -59.6) | 0.9 | (0.5 – 1.2) | -75.8 | (-85.7 – -68.7) | 0.2 | (0.1 – 0.7) |
| 70% | 0.3 | (0.1 – 0.5) | -91.1 | (-95.6 – -86.2) | 0.3 | (0.1 – 0.5) | -92.8 | (-97.2 – -88.0) | 0.1 | (0.0 – 0.2) |
| 90% | 0.1 | (0.0 – 0.1) | -98.3 | (-99.5 – -96.4) | 0.1 | (0.0 – 0.1) | -98.5 | (-99.6 – -96.8) | 0.0 | (0.0 – 0.0) |
| *Resistant strain less fit\*\** |  |  |  |  |  |  |  |  |  |  |
| 0% | 4.1 | (3.1 – 23.6) | -1.1 | (-27.5 – -0.9) | 4.0 | (0.1 – 10.2) | -2.6 | (-90.0 – 1.0) | 0.1 | (0.0 – 13.4) |
| 30% | 3.3 | (2.1 – 23.5) | -22.2 | (-40.6 – -6.3) | 3.1 | (0.1 – 10.2) | -23.3 | (-91.2 – 0.5) | 0.1 | (0.0 – 13.3) |
| 50% | 2.6 | (1.5 – 23.4) | -37.7 | (-52.9 – -6.8) | 2.5 | (0.1 – 10.1) | -38.5 | (-92.3 – 0.1) | 0.1 | (0.0 – 13.2) |
| 70% | 2.1 | (1.0 – 23.3) | -48.8 | (-67.4 – -7.2) | 2.1 | (0.1 – 10.1) | -49.7 | (-93.2 – -0.3) | 0.0 | (0.0 – 13.2) |
| 90% | 1.8 | (0.7 – 23.2) | -57.2 | (-76.8 – -7.5) | 1.7 | (0.1 – 10.1) | -58.0 | (-94.0 – -0.6) | 0.0 | (0.0 – 13.1) |

\* CrI: 95% credible interval of 343 runs.
\*\* 5% reduction in transmission probability

The estimated vaccination impacts were similar when assuming that selection for resistance can only occur upon treatment or only during the infectious period. However, assuming that resistance can occur during the infectious period only resulted in higher AMR prevalence in 2050 as every infected individual can develop AMR, compared to only a certain percentage of the population (the treated individuals) in the AMR upon treatment only scenario.

### Mechanism of vaccine action

Results were very similar if the vaccine reduces transmissibility, rather than susceptibility (Supplementary Tables S4.1). However, a vaccine that reduces the duration of infection is less effective: overall prevalence decreases less than with a vaccine that reduces susceptibility or transmissibility, and vaccination hardly delays the time to AMR development (Supplementary Table S4.2). An all-or-nothing vaccine is most effective of all vaccine types studied in reducing population prevalence (Supplementary Table S4.3). When vaccination uptake is high enough (>80%), AMR can be prevented even with low values of VE. When uptake is lower (40%), AMR development can only be prevented with a VE higher than 70%.

## DISCUSSION

In this mathematical modelling study, we showed that a vaccine that induces partial protection can prevent the development of AMR and reduce gonorrhoea prevalence, but only for high levels of vaccine effectiveness and high uptake in high sexual activity MSM, or if the duration of protection is long. A vaccine with characteristics comparable to the MenZB vaccine (i.e. 30% VE and short lasting immunity) is unable to prevent the development of AMR, but does increase the time to AMR development. An all-or-nothing vaccine would be most effective in reducing population prevalence and preventing AMR.

A strength of our mathematical model is the incorporation of AMR as a stepwise reduction in the proportion of fully sensitive strains that resembles the gradual drift towards observed increasing MIC values and decreasing susceptibility to ceftriaxone.^3,4,6^ Additional strengths are the use of empirical data about sexual behavior and gonorrhoea prevalence in Dutch MSM. There are also limitations to the model. First, the process of AMR development is much simplified. Given the incomplete empirical understanding of how the accumulation of resistance mutations result in ceftriaxone treatment failure,^23^ fitting the model to the distribution of MIC values over time was appropriate. There was a slight discrepancy between the model and data for the intermediate sensitivity MIC values (MIC >0.016 and <=0.064). In the GRAS data, an increasing trend was only seen since 2017, while in the model, a very slowly increasing trend for all years was observed. However, this increasing trend is in line with many countries that observe MIC drift.^3,4^

A second limitation is that we used a deterministic approach, despite the stochastic nature of AMR. This might influence the timing of AMR development, since stochastic events leading to extinction of the resistant strain are not taken into account. However, our results are robust for when resistant strains are frequent enough to evade stochastic extinction. Third, we combined the urogenital, anorectal and pharyngeal anatomical locations and averaged the percentage symptomatic infections and duration of infection. Incorporating each anatomical site separately would complicate the model parameterization substantially, introducing many parameters whose values remain unknown (infection, transmission and behavioural parameters). Fourth, we assumed vaccination of susceptibles only. This is probably a realistic scenario because high risk MSM are regularly tested, and vaccination, if ever rolled out, would probably be linked to testing before vaccination. Last, we assumed that, if a resistant strain is circulating, antimicrobial susceptibility testing would continue to be used for surveillance purposes only. However, this is likely to change resulting in a lower VE needed to curb AMR development.

Our results were less optimistic than another study that investigated the impact of a vaccine with characteristics comparable to the MenZB vaccine (i.e. VE of 30%) given to MSM.^25^ Whittles and colleagues found that vaccination could reduce gonorrhoea incidence by 45% to 75% by 2030, depending on the percentage of resistant gonorrhoea cases being treatable. Whittles et al. assumed outside introduction of a resistant strain, for which survival depended on the network into which it was introduced. Import of AMR *N. gonorrhoeae* is well-documented, including that of genotype 1407, which spread internationally, but disappeared owing to clonal replacement. ^S33^ In our study, *N. gonorrhoeae* resistance developed within the model population itself. This is a more realistic scenario if *N. gonorrhoeae* resistance mutations are acquired from commensal Neisseria species. These events could happen in several people at the same time, making it more difficult to curb transmission of AMR.

The mode of vaccine action of the MenZB vaccine and of future gonorrhoea vaccines is not known. We showed that a vaccine that reduced susceptibility had similar impact in reducing population prevalence as a vaccine that reduced transmissibility. A vaccine reducing the duration of infection was least effective. This finding probably results from the relatively high transmission probability per sex act for gonorrhoea; reducing the duration of infection does not prevent much transmission because transmission already occurred before clearance. An all-or-nothing vaccine had the largest impact on reducing population prevalence because it prevents infection and thereby the development of resistance. This illustrates the importance of investigating the exact mode of action in animal models and clinical trials in order to estimate the potential impact of a vaccine and the optimum vaccination strategies.

We showed that vaccination delayed the time to AMR development in *N. gonorrhoeae*. However, we do not know how much exactly, since resistance can develop quicker depending on mutation probabilities and the percentage treatment failure when infected with the resistant strain. Nonetheless, delaying the development of AMR is a crucial goal for the control of gonorrhoea globally,^1^ in view of decreasing susceptibility to ceftriaxone,^3^ the increasing emergence of multidrug resistant and extensively drug resistant strains^19-22^ and the limited alternative treatment options.^23^ There are currently promising new drugs in different stages of clinical development, such as zoliflodacin,^S34^ which could potentially be used in the future, if licensed. Besides vaccination, there are other ways in which AMR spread could be delayed, for example by the development of point-of-care tests that also detect determinants of antimicrobial susceptibility.^S35^ But the pipeline for molecular tests that detect resistance to extended spectrum cephalosporins is even less developed than for new antimicrobials.^S36^ In the absence of such tests, traditional culture and antimicrobial susceptibility testing should be expanded to inform national and global treatment guidelines.^S37^ If a resistant *N. gonorrhoeae* strain is circulating, the use of susceptibility testing will also be increasingly used to determine correct treatment, so if vaccination is part of a prevention strategy amongst MSM, a higher VE would be needed to curb AMR.

Vaccine development should be prioritized for pathogens for which AMR is an important or urgent threat. A randomized controlled trial of a new generation meningococcal group B vaccine (4CMenB (Bexsero) amongst MSM in the USA, will provide additional data on the impact of non-gonorrhoea specific vaccination.^S38^ Vaccines that prevents *N. gonorrhoeae* infection with limited efficacy, could delay AMR development in MSM, thereby providing additional time for the development of new antimicrobials and more efficacious vaccines.

## Data Availability

Data is available upon request

## Acknowledgements

We would like to thank Henry de Vries for useful discussions and Amy Matser for critically reviewing the manuscript. The Amsterdam Cohort Studies on HIV infection, a collaboration between the Public Health Service of Amsterdam, the Amsterdam University Medical Centers location AMC, Sanquin Blood Supply Foundation, MC Jan van Goyen and DC Clinics Lairesse, are part of the Netherlands HIV Monitoring Foundation and financially supported by the Netherlands National Institute for Public Health and the Environment. The ACS gratefully acknowledge all the study participants for their co-operation and participation, research nurses for collecting the data (Samantha de Graaf and Leeann Storey). The authors also thank Dominique Loomans, Ertan Ersan, Maartje Dijkstra, Liza Coyer, and Ward van Bilsen for data management.

## Author contributions

JH designed the study, developed the mathematical model, performed the model analyses and drafted the manuscript. NL contributed to the design of the study and NL, MV, MX to the design of the mathematical model. MB analysed the Amsterdam Cohort Studies data. All authors contributed to the interpretation of the results and commented on the manuscript.

